# Towards a framework for the scale-up of rehabilitation for patients with non-communicable disease in low-resource settings

**DOI:** 10.1101/2022.08.03.22278360

**Authors:** Martin Heine, Wayne Derman, Susan Hanekom

**Affiliations:** Stellenbosch University, Faculty of Medicine and Health Sciences, Institute of Sport and Exercise Medicine, Department of Sport Science, Cape Town, South Africa; Julius Global Health, Julius Center for Health Sciences and Primary Care, University Medical Center Utrecht, Utrecht University, Utrecht, the Netherlands; Stellenbosch University, Faculty of Medicine and Health Sciences, Division of Physiotherapy, Cape Town, South Africa; IOC Research Centre, South Africa

## Abstract

**Objective:** To explore and synthesize critical factors for the scale-up of comprehensive rehabilitation care for people with non-communicable disease in low-resourced settings.

**Methods:** A core set of 81 articles were selected from two published scoping reviews. Using the principles of thematic analysis, the core set was analysed for factors that may directly or indirectly affect the feasibility or scale-up of rehabilitation. Categories and themes were formulated via an iterative team approach using the core set (n=81). Subsequently, we triangulated the thematic analysis against our findings from a feasibility study conducted in a low-resourced, urban, setting in South Africa. Next, a validation article set (n=63) was identified by updating the searches for the respective published reviews, and a purposeful sample of articles drawn from the validation set (n=13; 20%) was used to validate the factors identified in the primary analysis based on the principles of data saturation.

**Findings:** A total of 40 different themes (i.e., critical factors) were derived from 169 categories. Subsequently the identified factors could be packaged into nine system elements principal for the scale-up of rehabilitation for people with NCDs in terms of increasing population coverage, increasing comprehensiveness, and integration within existing health structures.

**Conclusion:** A multitude of factors which affect the feasibility and scale-up of rehabilitation for NCDs in low resource settings were identified. These factors are multi-dimensional and multi-directional. Researchers and policy makers should consider these factors and their interconnectedness when planning to address the rehabilitation needs through implementation and scale-up initiatives.

## Introduction

Rehabilitation is embraced within the universal health coverage (UHC) target of the sustainable development goals as an essential health service, and moreover, access to rehabilitation is considered a human right.(1) Globally, the need for rehabilitation has been increasing; particularly, in low-to-middle income countries (LMIC), low-resourced settings, or vulnerable populations.(2–4) For many LMIC, one could argue that the colliding epidemics of both communicable and non-communicable disease is further stressing the rehabilitation needs,(5) and the gap between access and availability of comprehensive rehabilitation is increasing.(6)

Rehabilitation for people with non-communicable disease is often multi-component (e.g., exercise, education, nutrition) and inter-disciplinary, aimed at addressing underlying risk factors (e.g., physical inactivity) while optimizing function, activity, participation, and health-related quality of life.(7) The magnitude of the rehabilitation need is most evident from research conducted for patients with cardiac conditions. A global audit of cardiac rehabilitation programs revealed that in only 40% of LMIC such rehabilitation services are available; roughly translating to one cardiac rehabilitation spot available for every 66 patients with ischaemic heart disease.(6,8) However, this gap in the availability of cardiac rehabilitation specifically, is illustrative for a broader need for rehabilitation type services in general, including rehabilitation for lung health,(10) access to assistive technology,(11) cardiometabolic disease, and others.(12,13) To address these needs, a concerted effort is needed to scale-up access to rehabilitation services (Figure 1).

**Figure 1:**
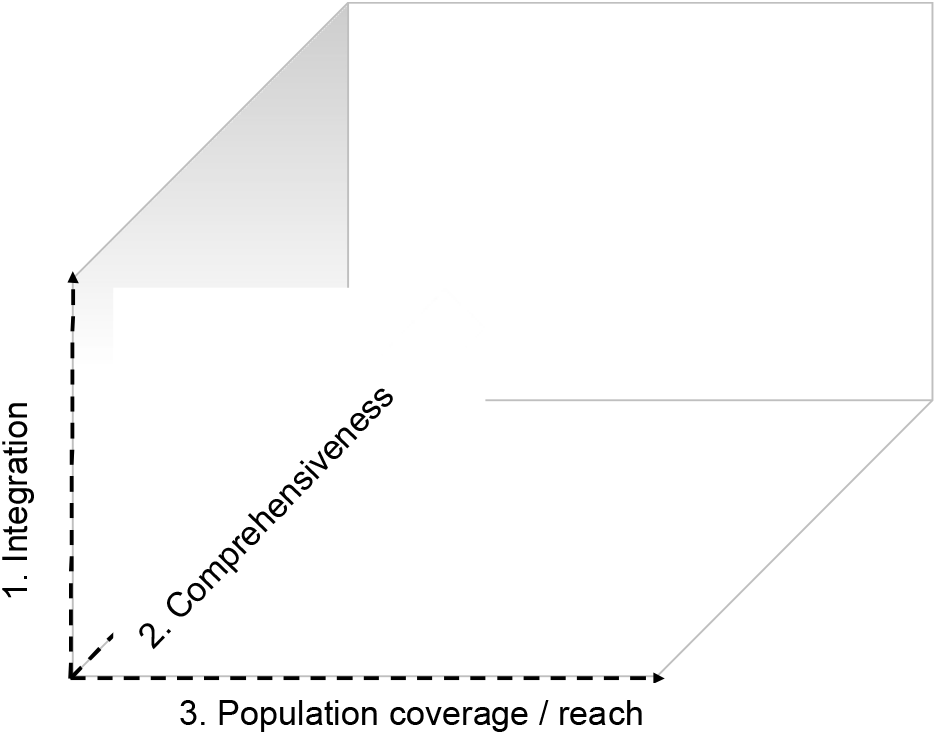
Three dimensional framework for the scale-up of integrated care adapted for rehabilitation: 1) strengthening integration within primary care structures including integrating rehabilitation care for NCDs within existing programs for communicable disease management (e.g. integrated care for Diabetes in patients with HIV/AIDS); (2) increasing the comprehensive nature of existing programs to ensure more core components of rehabilitation for NCDs are included; and (3) set-up of new programs or program optimization to increase the number of people that have access to rehabilitation services (based upon van Olmen et al. 2021(14)).

Scale-up is defined as the ‘efforts to increase the impact of health interventions so as to benefit more people and to foster policy and programme development on a sustainable basis’.(15) We’ve adapted the scale-up framework for an integrated care package,(14) to a three-dimensional scale-up framework that applies to rehabilitation: (1) strengthening integration within primary care structures including integrating rehabilitation care for NCDs within existing programs for communicable disease management (e.g. integrated care for Diabetes in patients with HIV/AIDS); (2) increasing the comprehensive nature of existing programs to ensure more core components of rehabilitation for NCDs are included; and (3) set-up of new programs or program optimization to increase the number of people that have access to rehabilitation services (Figure 1). The on-the-ground-realities in low-resource settings,(3) add real layers of complexity to the scale-up of rehabilitation as to mitigate the burden of NCDs.

Therefore, the objective of this study was to (i) review critical factors that affect the feasibility and scale-up of rehabilitation care within the NCD population, (ii) triangulate those findings with our lived experiences in a South African low-resourced setting, and (iii) reflect on how these critical factors may inform scale-up of rehabilitation and address the implementation gap.

## Methods

### Data sources

For this study, we used two data sources.

#### Existing body of evidence (source 1a and 1b)

First, we included articles pertaining to the rehabilitation of people with non-communicable disease in low-resource settings (i.e., the core article set). These articles were derived from two previous reviews, conducted by members of our team, within the NCD space,(3,4) The two reviews combined, comprised 108 unique articles pertaining to either i) exercise-based rehabilitation for people with NCDs living in low-resource settings, or ii) studies in the field of rehabilitation, making reference to the research being conducted in a “low-resource setting”.(3,4) From the 108 unique articles, a total of 81 studies specifically included an NCD population and hence these 81 articles were included for thematic analyses to identify factors that affect the feasibility and scale-up of rehabilitation for people with NCDs in low-resource settings (source 1a). Subsequently, we updated both searches underlying these two published reviews, with a specific focus on studies conducted in an NCD population (see Figure 2). The updating of these two reviews led to a second set of articles (source 1b), in addition to the 81 studies already identified, that we used for validation of the initial thematic analysis (i.e., validation set; n = 63).

**Figure 2:**
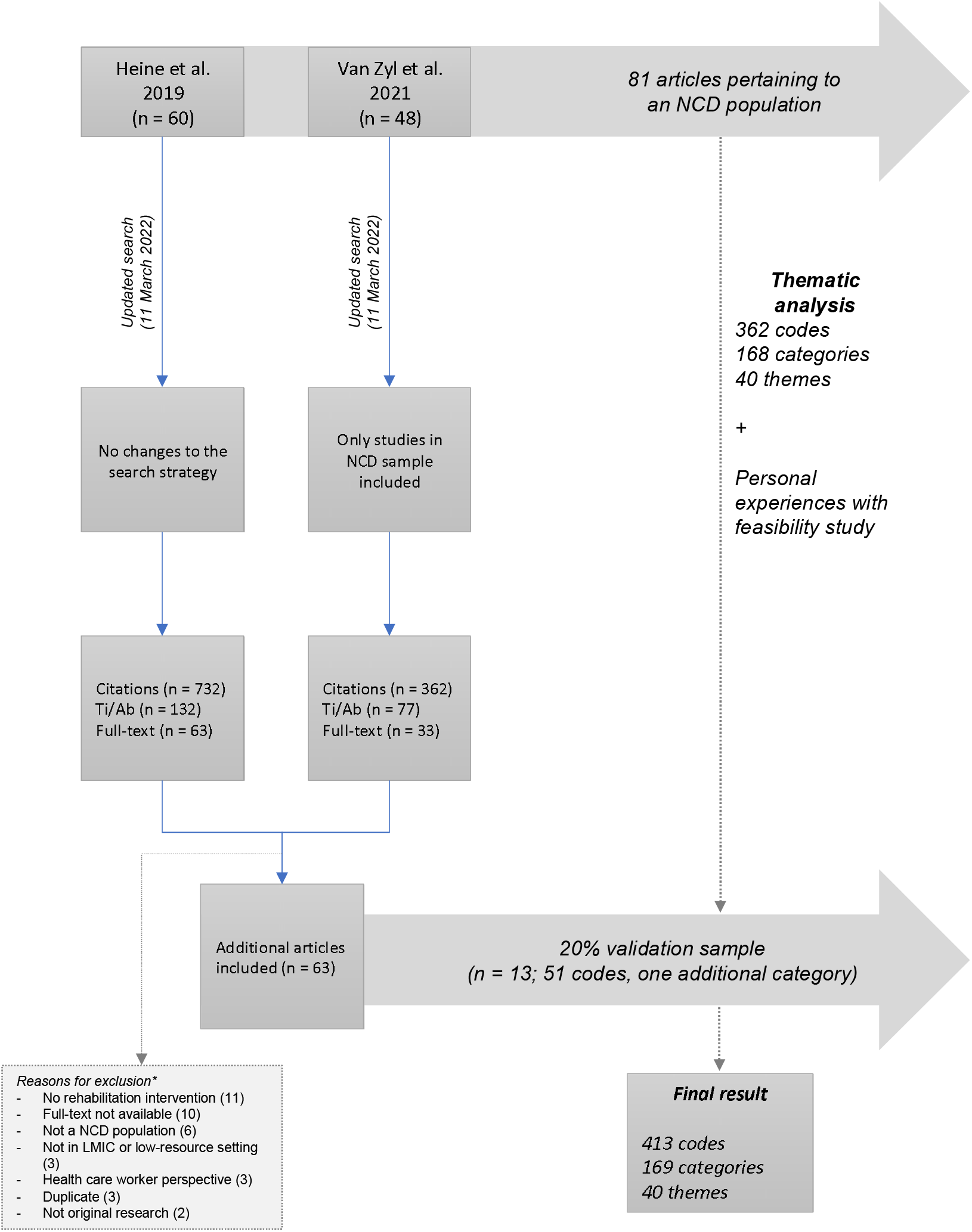
Study selection flowchart. *multiple reasons per excluded study possible.

#### Primary research (source 2)

The second data source comprised our own experiences in conducting a feasibility randomized clinical trial of a patient-centred rehabilitation program for patients with non-communicable disease (n=74) living in a South African low-resource setting.(16–18) For this primary research, ethical approval was obtained from the Stellenbosch University Health Research and Ethics council (HREC2-2018-0913), and Western Cape Health Research Committee (WC-201806-027). The trial was prospectively registered (PACTR201807847711940). All participants provided oral and written informed consent. This study took place within a public sector, primary care community day centre (CDC) and adjacent rehabilitation unit (Cape Town, South Africa). The specific community is one of many historically displaced communities in Cape Town: an urban settlement of >54,000 residents battling adverse social determinants of health, including (extreme) poverty, unemployment, substance abuse, gangsterism, trauma, and limited access to (quality) education.(19) The rehabilitation unit is a service-learning site,(20) providing a variety of rehabilitation services through the placement of undergraduate students in training under supervision of government-funded university lecturing staff. Patients offered the program could make an informed decision to participate (similar to decision making within routine clinical practice).(17)

### Thematic analysis

A thematic analysis was conducted by coding the 81 articles (i.e., source 1a) for direct and indirect factors that may influence the feasibility or scale-up of rehabilitation for NCDs in a low-resourced setting. Coding took place both inductively and deductively, guided by the socio-ecological model.(3,21) The socio-ecological model is particularly useful when determinants of an outcome are nested within different levels of influence.(22) Once a code was identified, we did not attempt to code subsequent articles using the same coding as we argue that how *often* a code was applicable does not necessarily equate to its relative importance to the wider system. In other words, a code could potentially occur only once while having more “weight” than a code that transpires numerous times. Once all articles were coded, we merged all codes into content categories and themes (i.e., key actors) through a series of team meetings. The identified key factors were subsequently triangulated against our own experiences (source 2). Any actors not identified via thematic analysis of secondary research, which were found especially relevant in our primary research, were added.

### Validation

Subsequently, we used a data saturation process to validate the themes derived from the thematic analysis of primary (i.e., source 2) and secondary data (i.e., source 1a). This process is similar as for qualitative interview-based research, where one seeks to reach saturation (i.e., no new categories or themes emerge). We decided *apriori* on a 20% sample from the validation set (i.e., source 1b) to reach data saturation. If any additional themes emerged from this validation step, a second 20% articles were selected, until complete saturation was reached. We used two processes to select articles for this validation step. First, we used purposeful sampling to select articles underrepresented in the core article set (source 1a), including underrepresented settings (i.e., low-income countries) and patient populations (i.e., cancer). Subsequently, the purposeful selection of studies from low-income countries, or in cancer patients, was supplemented with a random selection of articles from the validation set until the 20% target was reached.

### Towards a framework for scale-up

Finally, we developed a preliminary conceptual synthesis of the themes derived from the thematic analysis through iterative team discussion. This additional synthesis aimed to identify common system elements that may enable scale-up and optimization of secondary prevention programs for NCDs in low-resourced settings, further condense some of the complexity, and provide a conceptual starting point in better understanding the system requirements for the scale-up of secondary prevention for NCDs in low-resourced settings.

## Results

### Study selection for validation set

The searches for both reviews were updated (11th of March 2022). A custom flowchart is provided in Figure 2 to showcase the selection process. A total of 1094 articles were screened at title and abstract level by two researchers independently using CADIMA software.(23) Subsequently, 209 abstracts were screened against the eligibility criteria, leading to 96 full-text articles for consideration, of which 63 met all criteria. At this stage, reasons for exclusion included “no rehabilitation intervention” (n = 11), full-text not available (n = 8), no NCD patient sample (n = 6), only healthcare worker perspective provided (n = 3), or other (n = 5). Thirteen articles (20%) were selected from this validation set, based on being conducted in a low-income country (n = 5), patients with cancer (n = 4), or by random selection (n = 4). A full reference list (online supplement 1) and consolidated overview (online supplement 2) of all 144 articles (source 1a and 1b) can be found elsewhere.

### Thematic analysis

The initial thematic analysis resulted in 362 codes, drawn from 81 articles. Subsequently, these codes were merged into 168 categories, and 40 themes. In the validation step, based on 51 codes, one additional category was identified (i.e., seasonal resource fluctuations), yet no additional themes were formulated. As such, no second set of articles was selected from the validation set. The final analysis comprised 413 codes (362 plus 51), drawn from 94 articles (81 plus 13), leading to 169 categories and 40 themes; twenty-four of which we recognised from the case example. Based on the single new category identified, no second random selection of articles was made. A description of each theme can be found in online supplement 3. For each theme, the main spheres of influence, according to the socio-ecological model are provided (Figure 3). Finally, themes are clustered (Figure 3, inner circle) into sub-systems that directly or indirectly may inform program optimization to promote program integration, comprehensiveness and/or reach (Figure 4).

**Figure 3:**
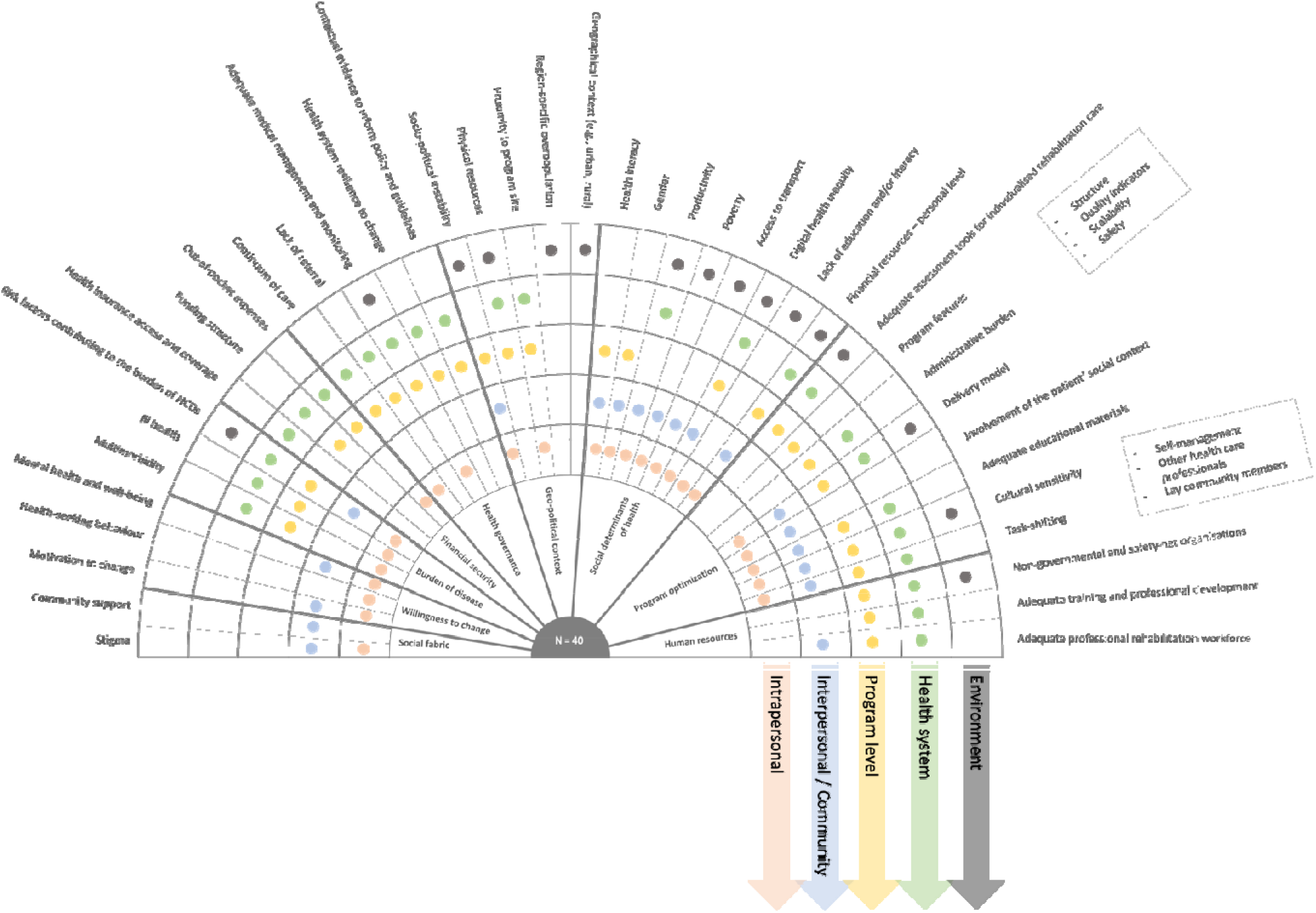
Overview of the 40 themes (outer circle) identified from 169 categories, their relation to the socio-ecological model (colour coded), and clustered according to nine different sub-systems (inner circle; Figure 4). A description of each theme can be found in online supplement 2.

**Figure 4:**
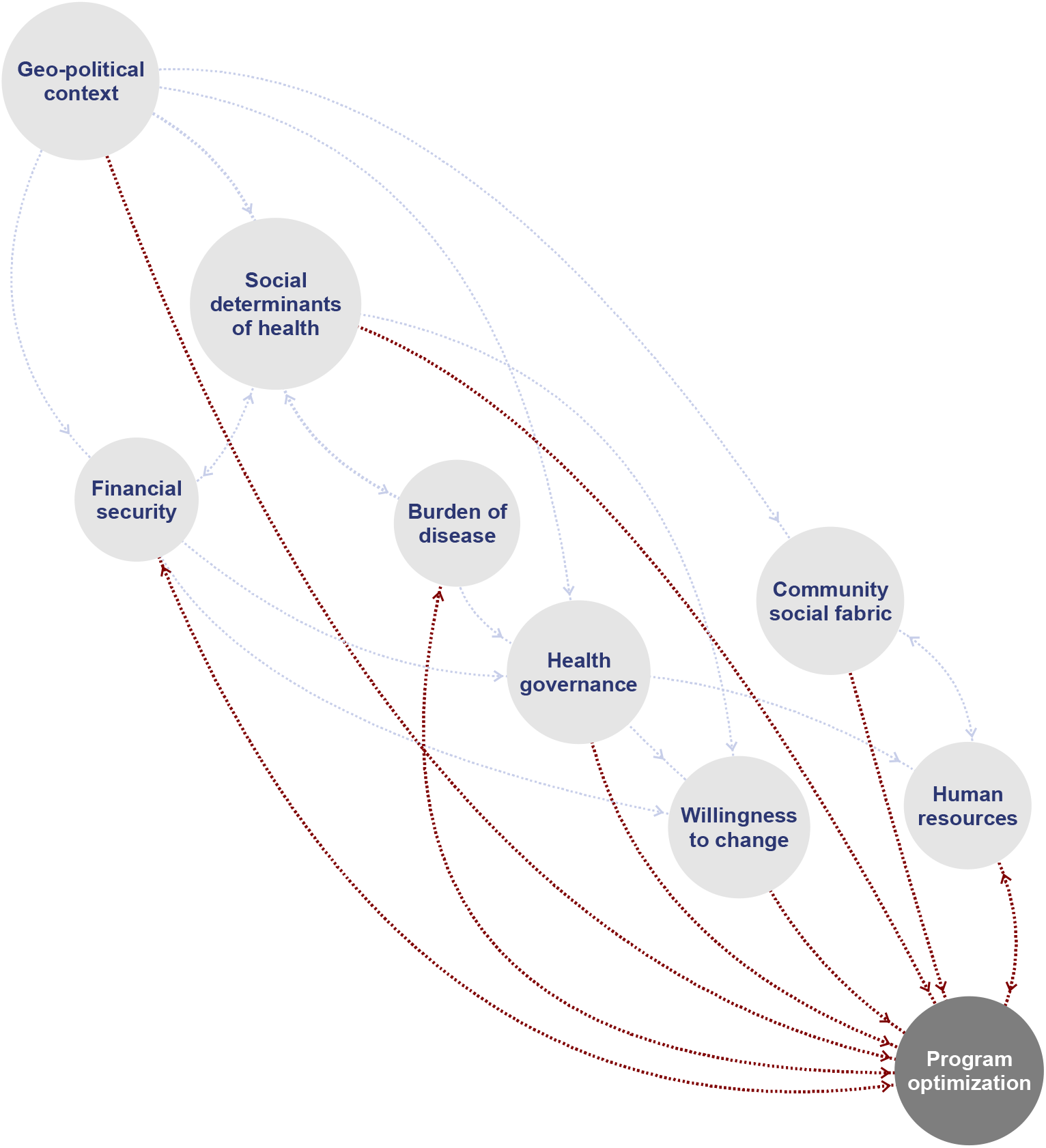
Systems map of mechanisms through which different sub-systems may directly (red) or indirectly (grey) inform program optimization, and as such promote the integration, reach, and comprehensiveness of rehabilitation for people with NCDs in low-resourced settings.

## Discussion

As the burden of NCDs is increasing globally, disproportionally affecting those in low-resourced settings, so is the parallel gap between the need and access to comprehensive rehabilitation.(12) To our knowledge, this is the first paper to approach the need for the scale-up of rehabilitation care for NCDs in a low-resourced settings from a holistic and complexity lens. This paper outlines 40 critical factors, drawn from multiple data sources, collated into 9 sub-systems, that may inform the scale-up and optimization of rehabilitation in low-resourced settings. We argue that these factors are imperative to the (i) integration of rehabilitation within in existing health structures, (ii) the comprehensive nature of the rehabilitation program offered, and when attended to adequately, will likely (iii) increase population coverage, reach, and access. The nine sub-systems (Figure 4) provide a template for future research and scale-up initiatives aimed at closing the rehabilitation need versus access gap. Herein, we reflect on the critical factors and sub-systems presented, in relation to the three dimensions of scale-up.

### Integrated Rehabilitation

The first dimension of scale-up is integration of care within existing health structures. Within the rehabilitation space, we could distinct three forms of “integration” that are particularly relevant to address the rehabilitation need. Firstly, integration of rehabilitation care for multiple conditions simultaneously to address multimorbidity or converging burdens of disease.(24) Secondly, integration of rehabilitation care with other social domains that therefore can target social determinants of health.(25) Thirdly, integration of rehabilitation within primary care.(26) The value of each of these three becomes apparent when reflecting on the critical factors and sub-systems identified in this review. In many low-resourced settings, multiple burdens of disease collide with human resources (i.e. quantity, adequate training) and health system governance unequipped to tackle the convergence of, for example, communicable and non-communicable disease or diabetes and hypertension as in our case example.(16,27–29) Hence, within rehabilitation care for NCDs too, we may need to move beyond disease-siloed programs (e.g., cardiac rehabilitation, pulmonary rehabilitation) towards person-centred care models of holistic - chronic disease rehabilitation and provide real-world evidence in their support.(18) In providing such support, key questions for rehabilitation in low-resourced settings remain with respect to feasibility, adequate evaluation to inform policy, and impactful implementation.(30)

Furthermore, there is clear evidence that factors outside of health care delivery greatly affect an individual’s health and well-being; this is particularly true for low-resourced settings with a high prevalence of adverse social determinants of health, and including rehabilitation.(31) In addition to the impact of social determinants of health on health and well-being, these determinants also directly impact the feasibility of and access to rehabilitation care. Some critical factors and social determinants of health identified in this mixed-methods review were for instance health literacy, health-seeking behaviour, poverty, productivity, and access to transport. Such social determinants of health may inform both the burden of disease through their association with underlying risk factors,(32) financial means (e.g., out-of-pocket expenses) and security (e.g., access to health insurance) to engage with rehabilitation services, and willingness or ability to make lifestyle changes that can contribute to risk reduction. Hence, rehabilitation-led models of integrated rehabilitation- and social care (e.g., case manager) that comprehensively address social determinants of health can make a remarkable difference in the burden of disability and health inequities.(25) Awareness of individuals‘ social needs, adjustment of care provided, assistance for the individual to have needs met, alignment of efforts with community and social care organizations, and advocacy for changes to infrastructure and policy (both health and social) may facilitate such rehabilitation-led models.(25)

Primary care is a model of care that supports first-contact, accessible, continuous, comprehensive, and coordinated person-focused care. The World Health organisation has specifically called for strengthening and scale-up of NCD interventions within primary care in their draft 2023-2030 roadmap on the prevention and control on NCDs.(33) Various health systems have approached the integration of rehabilitation into primary care differently, reflecting differences in the organisation of care and health governance.(34) Six different ways of integrating rehabilitation in primary care have been described, including clinic-based, outreach, self-management, community-based rehabilitation, shared care, and case management.(26) Each of which emerge as possible models in relation to the subs-systems identified in this review, including the geo-political context (e.g., rural context informing an outreach model), human resources (e.g., task-shifting, self-management), and health governance (e.g., resilient health systems, referral pathways). The work presented here, in conjunction with various models for integrating rehabilitation into primary care, including social care where applicable, and cognisant of multimorbidity, could inform program optimization and scale-up initiatives.

### Comprehensiveness

Integrated models of rehabilitation- and social care already speak to a more comprehensive offering of rehabilitation beyond those core components (e.g., physical inactivity, nutrition) strictly related to the underlying risk factors for non-communicable disease. In augmenting the comprehensiveness of rehabilitation, one could argue that human resources are central, including both quantity and quality (e.g., adequate training). In light of social determinants of health and integration within primary care, such human resources may not be restricted to allied health professionals like physiotherapist and also include non-medical human resources (e.g., community champions, community health workers, caregiver-led models). To optimize the human resources required to provide quality comprehensive rehabilitation for NCDs in low-resourced settings, one needs to ask a couple of questions. First, what structural risk factors (e.g., physical inactivity, nutrition, health illiteracy, poverty) are underlying the burden of disease in my specific context and how may that inform the human resources required (i.e., training needs, composition of the rehabilitation team, integration with social care)? Second, what non-medical human resources are available in my specific context, and how can we optimize program delivery by actively engaging these resources (e.g., task-shifting, self-management)? Importantly, the process of task-shifting in itself may be complex, with important roles for health literacy in self-management models,(35) and the community’ social fabric in the feasibility of training lay community workers.(36) Thirdly, how can digital innovation be used equitably to promote access, uptake and/ or adherence of technology driven rehabilitation models, and, how does this affect the competencies required of those overseeing such rehabilitation services? The implications thereof, for example, may be that a single physiotherapist oversees a cadre of community health workers for coordinated physiotherapy and exercise training using a digital telehealth platform rather than solely providing face to face clinical services. In this light, we argue that it is pivotal that while we train the next cadre of rehabilitation professionals,(37) we embrace the competencies needed to set-up and coordinate such task-shifting models as a means to optimize rehabilitation delivery.

### Increasing access

This review provides an overview of the various critical factors that may be considered in strengthening existing, and scale-up of new, rehabilitation programs. However, addressing a single factor is unlikely to result in the provision of quality rehabilitation care, as each factor should be recognised as equally important, yet equally inadequate if addressed in isolation. We argue that the critical factors, the nine sub-systems, in conjunction with existing frameworks (e.g., WHO rehabilitation competency framework) that relate to these sub-systems can assist research and policy makers in making informed decisions in the scale-up of rehabilitation, or in conceptualisation the research and information needs. What becomes apparent from this work, is that scale-up of rehabilitation is complex, and requires intersectoral and interdisciplinary impetus across the continuum of care.

### Limitations

The process and sources to identify critical factors which affect the feasibility of rehabilitation in low resource settings, can be considered objective and reproducible. However, one could argue that the process used that led to identifying sub-systems was one-dimensional (i.e., all team members are [allied] health professionals). Given the call for an intersectoral approach to address the rehabilitation need, involving broader stakeholder groups, which include patients, communities, and government agencies in identifying the interplay between factors could add to the richness and complexity. Subsequently, this may further enhance our understanding of the system within which rehabilitation is to be provided. However, such pre-existing “systems” will be highly context specific. Hence, we argue that this work should be considered a template of factors that can be considered in the scale-up rehabilitation and program optimization, yet that contextualisation to any specific situation is required.(38)

## Conclusions

The present study highlights the complexity of rehabilitation to people with NCD living in low-resource settings. The systems lens not only provided a framework and checklist to foster a rich understanding of the factors influencing the feasibility of rehabilitation in this unique context, but also highlighted the interdependence of the factors. The complexity identified, encourages innovative an intersectoral approaches to address the rehabilitation needs in these settings.

## Data Availability

All data produced from published literature are contained in the manuscript or supplementary files. Primary data is available upon reasonable request to the authors.

https://data.mendeley.com/datasets/fbhh8dmz7w

## Notes

### Competing Interest Statement

The authors have declared no competing interest.

### Clinical Trial

PACTR201807847711940

### Clinical Protocols

https://pubmed.ncbi.nlm.nih.gov/30975678/

### Funding Statement

Parts of this study were funded by the AXA Research Fund (S005949)

### Author Declarations

Ethical approval was obtained from the Stellenbosch University Health Research and Ethics council (HREC2-2018-0913), and Western Cape Health Research Committee (WC-201806-027).

