## Supplemental File 3 for "Towards a framework for the scale-up of rehabilitation for patients with non-communicable disease in low-resource settings"

### Online supplement 2: Overview of the 40 themes identified and their relation to the socio-ecological model.

**Access to transport.** Availability of personal transport (e.g., car), availability, affordability, and safety of public transport (e.g., bus, taxi).

**Adequate assessment tools for individualised rehabilitation care.** Tailoring the program content and structure to meet individual patient needs may be essential in achieving comprehensive health benefits. Individualised assessment tools that are simple and accessible that can inform individualised rehabilitation care are imperative.

**Adequate educational materials.** Educational materials are used to promote health literacy, health-seeking behaviour, or in the context of self-management. Adequate education material is personalised, provided in a manner conductive to the patient’ context (including when provided through e-solutions), and tailored in terms of cultural sensitivity, gender, and language. The information needs may shift throughout the continuum of care in relation to the chronicity of NCDs.

**Adequate medical management.** Detection of people at risk, routine monitoring, communication between health professionals through the continuum of care, and so forth. Inadequate medical management may affect the health and well-being of the participants, with implications for the, amongst others, referral to the rehabilitation team, multimorbidity, and program safety. In some settings, adequate medical management may only be available when insured or with significant out-of-pocket expenses.

**Adequate professional rehabilitation workforce.** Quantity and quality of human resources known to be important in the provision of (multi-disciplinary) rehabilitation, including allied health professions, medical oversight of (high risk) patients, administrative staff, psychologists, nurses, and others. In some settings, tasks are shifted to non-conventional health professionals, lay community members, or an increased focus on self-management.

**Adequate training and professional development.** Continued development of those involved in the rehabilitation process, includes knowledge, skills, and competencies of health professionals, but also adequate training when tasks are shifted to lay community members. The training needs may shift in relation the prevailing risk factors or disease burden.

**Administrative burden.** The time and effort required for program administration and record keeping. Administrative burden, for example, is increased in settings where patient information cannot be easily shared (e.g., paper records), or in settings where patients are difficult to reach (e.g., rapid changing contact details).

**Community support**. A community social fabric where people with non-communicable disease are socially supported. For instance, related to the stigmatisation of disease, and may influence mental health and well-being.

**Contextual evidence to inform policy and guidelines.** High-quality evidence for the cost benefits of rehabilitation in a specific setting can assist in developing context specific guidelines and policy. The “benefits” need to align with both the patient-perspective and key information required for upscaling and policy initiatives. Protocolised care may improve quality and consistency in rehabilitation program’ offering and perceived value.

**Continuum of care.** Non-communicable disease is inherently chronic, and therefore poses important challenges to provide long-term adequate care across the continuum.

**Cultural sensitivity.** Feasibility of rehabilitation is partly dependent on how well the program is aligned with cultural and / or religious features common to the catchment area. For example, in some settings, programs may need to be gender specific to remove culturally informed barriers to participation. In other settings, the type of exercise therapy needs to be adjusted to the local context or educational materials need to be developed cognisant of cultural, religious, or language-related factors.

**Delivery model.** Inpatient, Outpatient, home-based, telehealth, hybrid, out-reach were some of the delivery models identified. The delivery model most adequate, may depend amongst others, on the desired program structure, patient population, program safety, geographical context, human resources (including task shifting), funding structure, and digital health equity.

**Digital Health Equity.** Digital health innovation needs to be scalable and accessible to the local demographic and may mean that the latest technological advances are excluded. Challenges may relate to access to information technology, digital literacy, and personal financial resources.

**Financial resources – personal level**. Rehabilitation has the potential to alleviate the impact of disease in financial well-being. Conversely, in absence of social security measures, work was an important barrier to rehabilitation uptake and adherence. Personal financial resources, in relation to rehabilitation, may also affect the ability to pay for health insurance, out-of-pocket expenses, and transport. Moreover, financial resources may vary with time (e.g., more income during specific seasons) and therefore may affect continuity of care.

**Funding structure.** The way (e.g., government, private) the provision of rehabilitation is funded may affect access, human resources, scalability, amongst others.

**Gender.** There are significant differences in the barriers experiences to participate in rehabilitation between men and women. For example, there may be cultural / religious barriers that would require women-specific programs, the disease burden and comorbidity profiles may differ, or family responsibilities may hamper participation. Conversely, in some settings, men may be primarily responsible for income generation which would argue for rehabilitation, yet also poses an important barrier to participate.

**Geographical context.** Urban and rural are the two most common denotations that speak to the geographic context. Low-resourced settings can be found in both, however, with different features that impact the feasibility of rehabilitation. For example, in a rural context, proximity to health care providers may be challenging and hence out-reach programs may be more conducive. Conversely, out-reach programs can be challenging in relation to the chronicity of NCDs. In an urban context, challenges may be related to community safety or violence, to overpopulation, or overburdened public health facilities.

**Health insurance.** Affordable health insurance with adequate coverage with increase the feasibility of rehabilitation.

**Health literacy.** Various aspects of health literacy influence the feasibility of rehabilitation through various mechanisms, including health-seeking behaviour. However, health literacy also informs program requirements, education materials, ill health and multimorbidity. Various social determinants of health (e.g., education, poverty) will affect health literacy levels.

**Health systems resilience to change.** The burden of disease in many low-to-middle income countries is shifting from communicable disease to non-communicable disease. The convergence of the two poses a significant threat to health systems that are less resilient to adapt to the different care requirements, which can be exemplified by inadequate funding allocation, professional development, and human resources.

**Health-seeking behaviour.** Closely related to health literacy and motivation to change is health-seeking behaviour as the active pursuit of care when needed.

**Ill health.** Uncontrolled hypertension and blood glucose are commonly indicated as reasons for non-adherence or barriers to rehabilitation uptake. Imposes a need for thorough risk screening, stratification, and medical oversight prior to and during exercise-based rehabilitation. May impact program safety and needs for medical oversight and ability of the health system to respond to adverse events.

**Involving the patient’s social context.** Programs in which family-members join the treatment are considered more enjoyable and motivating. Closely related to social support.

**Lack of referral.** In many settings, referral from the tertiary care facility, to either primary or secondary care rehabilitation programs is inconsistent or absent. In absence of adequate referral structures, the ownership for continued care is shifted to the patient and therefore the uptake of rehabilitation increasingly dependent on health literacy and health seeking behaviour – both affected by a series of social determinants prevalent in low-resourced settings. Information technology may reduce the administrative burden associated with referral where possible.

**Level of education and/or literacy.** Quality education may be one of the key social determinants of health. In absence of quality education, there are considerable implications to the feasibility of rehabilitation, including health literacy, digital health literacy, and impact on program-specific or educational materials.

**Mental health & well-being.** Poverty, food or financial insecurity, socio-economic instability ill health and other all may affect mental health and well-being. Indirectly, these factors may affect the uptake and adherence to rehabilitation, for instance through reduced motivation to change.

**Motivation to change.** The insight and motivation that a change is required to improve health. With respect to rehabilitation of NCDs, this is important in for risk factor modification including physical inactivity, substance use, and nutrition. Such motivation may be expedited in a context where change is encouraged and facilitated.

**Multimorbidity.** The convergency of maternity related disease, trauma, communicable and non-communicable disease, in conjunction with inadequate medical care, and adverse social determinants of health implies a high level of multimorbidity in low-resourced settings. This affects various aspects related to the feasibility of rehabilitation in these settings, amongst others, the scale of rehabilitation needed, access to the program, complexity of patients (and subsequent knowledge and competencies of the program), program structure and delivery model.

**Non-governmental or safety-net organisations.** In some settings, rehabilitation is provided through non-governmental (e.g., out-reach programs) or safety-net organisations (e.g., service-learning driven programs). The organisations address important service gaps.

**Out-of-pocket expenses.** Monetary expenses associated with receiving care, including over the counter medication, but also indirect costs like transport.

**Physical resources.** Physical infrastructure (e.g., buildings) and equipment needed to provide adequate rehabilitation to patients with NCDs. Factors that may affect physical resources, amongst others, are the geographical context (e.g., rural), funding structure, socio-political stability.

**Poverty.** Lacking enough resources to provide the necessities of life, including access to health care and education.

**Productivity.** Beyond financial resources, productivity also entails the ability to participate in meaningful life-roles including family responsibilities, household tasks, social responsibilities, and volunteer work.

**Program features.** There are multiple factors that relate to adequate rehabilitation programs in low-resourced settings that pertain to how these *programs are structured* (e.g., number of sessions, duration). In part, this may be related to the available resources, including ability to shift task, and inform the delivery model. Program structure, disease population, medical management, available human resources, and delivery model may inform *program safety* and *scalability****.*** *Program quality* can be enhanced by ensuring adequate effective communication structures, safety protocols, as well as structured reporting and monitoring.

**Proximity to program site.** Distance between the patient with NCDs and nearest rehabilitation program. Can be bridged using task-shifting type rehabilitation models or using eHealth solutions.

**Region-specific overpopulation.** Particularly in urban low-resourced settings, there may be a considerable mismatch between health care / rehabilitation needs and health care / rehabilitation resources. In some settings (e.g., townships) such needs tend to cluster parallel to poverty, adverse social determinants of health, socio-political instability, and others.

**Risk factors contributing to the burden of NCDs.** In different settings different risk factors may be more prevalent. This will affect the type of NCD burden, yet may also affect program composition, required human and physical resources, feasible task-shifting, skills, and competencies.

**Socio-political instability.** The socio-political dynamics in some settings can be volatile, including protest action or gang violence amongst others. These dynamics may impact the feasibility of rehabilitation (e.g., access, human resources, safety), as well as the need for rehabilitation programs to remain amphibious to a level of uncertainty at a personal and/or community level.

**Stigma.** A setting in which disease, or (components of) rehabilitation are viewed in a negative way. For example, in some settings women are not expected to participate in moderate to vigorous intensity leisure exercise.

**Task-shifting.** Considering (human) resource constraints, various forms of task shifting emerged from the literature. Firstly, rehabilitation programs strongly focussed on self-management. Secondly, tasks conventionally attributed to allied health professionals were shifted to, for example, nursing staff, pharmacists, or other health professionals more likely to be available. Thirdly, there is also a considerable drive training and leveraging the use of lay community members for the provision or support of rehabilitation, including peer-educators, community health workers, or care navigators.
